# Hammersmith Neonatal Neurological Examination of healthy term infants at ages 6 and 10 weeks in Tshwane District, South Africa

**DOI:** 10.1101/2022.07.08.22276184

**Authors:** Marna Nel, Ute Feucht, Helen Mulol, Carina Eksteen

## Abstract

**Background:** The neurodevelopmental progress of infants below 3 months is globally not well described. A lack of published data on normative values of child development in this age group has been identified. In South Africa (SA), neurodevelopmental characteristics of infants at well-baby 6- and 10-week visits are omitted from the ‘Road to Health Booklet’, a nationally used patient-held clinical record. The neurodevelopmental status of infants in these age groups is not routinely monitored and data not documented. Important changes occur in the maturation of the central nervous system of infants at 6 weeks that mark this age as an important milestone for monitoring neurodevelopmental progress.

**Methodology and findings:** A prospective longitudinal study was performed on a sample of 35 healthy term-born, infants from low-risk pregnancies at 6- and 10-weeks postnatal age in the Tshwane District, SA. The status of infants’ posture, tone, reflexes, movements, orientation and behaviour were recorded on the Hammersmith Neonatal Neurological Examination (HNNE). Cut-off points on the 5th and 10th centiles, according to the HNNE ‘optimality scoring system’, were applied to the raw scores of the 34 items in the proforma evaluation form. This study quantitatively supports patterns of characteristic change occurring in muscle tone, posture, and visual behaviour of infants between 6 and 10 weeks.

**Conclusion and significance:** This study recorded data on the neurodevelopmental assessment of infants from low-risk pregnancies at 6- and 10 weeks post-term age in Tshwane District, SA. The optimality scores obtained in this initial study in a low-middle-income country can guide health professionals using this method of examination during early neurodevelopmental screening at well-baby clinics. Further research is necessary to develop a SA norm for identifying the motor and neuro-behavioural characteristics of 6- and 10-week-old infants.

## Introduction

Neurodevelopmental progress in early infancy (age below 3 months) is globally not well described. Adverse maternal and foetal factors leading to stillbirth e.g., placental insufficiency, often cause morbidities such as foetal growth restriction, premature birth, and neonatal encephalopathy in surviving and high-risk infants [1–7]. These conditions are associated with insults to the foetal and neonatal brain and result in altered neural pathways from an early age leading to deviant neonatal behaviour, impaired neurological maturation, and neurodevelopmental delays [8–15].

Developing countries are burdened with high numbers of young children who fail to reach their developmental potential [16, 17]. Hence, health care professionals are urgently called to identify at-risk infants and contribute to sustainable early childhood intervention by addressing and limiting adverse short- and long-term development sequelae in surviving and high-risk infants [7, 14, 16, 18–25].

The development of infants beyond the neonatal period, in particular at the 6- and 10-week postnatal age intervals, has been found to lack standardised normative values and detailed descriptions of milestone characteristics. In a follow-up study of healthy term infants, Guzzetta et al., (2005) described changes that occurred between birth and 3- to 10 weeks post-term age in the domains of *muscle tone, posture*, and *visual orientation*. The characteristics of change identified in these domains emphasized that the 6 weeks post-term age provides notable information and guidance that may enable health care professionals to identify and document the neurological maturation of infants beyond the neonatal period [26].

In South Africa (SA) health care professionals use the patient-held clinical record called the ‘Road-to-Health booklet’ (RtHB) to document child growth, immunization and developmental progress during routine postnatal follow-up visits at primary healthcare facilities [27]. This RtHB is incomplete in its description of developmental milestones for infants aged 6 and 10 weeks. The public health sector in SA provides services to approximately 84% of its 55 million people and the current birth rate per woman is 2.4% [28]. This means that a great number of infants in the age groups 6 and 10 weeks are not routinely monitored in terms of their neurodevelopmental progress and data regarding their neurological maturation process is therefore not documented [29].

The specific characteristics in the RtHB that describe the neurodevelopment in the domains of *hearing/communication, vision/visual adaptation, cognition/behaviour, motor skills*, and *the measurement of head circumference* for 6- and 10-week-old term infants have not yet been included in this RtHB [27].

The repertoire of neurodevelopmental characteristics omitted from the RtHB correlates with the domains of infant neurodevelopment contained in the Hammersmith Neonatal Neurological Examination (HNNE) [27, 30]. Dubowitz et al., (1980) developed the HNNE in which the fundamental characteristics in the domains of *posture and tone, tone patterns, reflexes, movements, abnormal signs* and *orientation and behaviour* of newborn term infant development are illustrated and described [30, 31]. The tool’s ‘*optimality scoring system’* enabled a quantitative association of prenatal motor activity with optimal scores in the domains of reflexes and movement patterns in newborn infants from low-risk pregnancies and predicted neurodevelopmental disability at one year [9, 32]. Normative data based on the HNNE optimality scoring system is available for preterm infants at term equivalent-, late preterm-, and infants at term age [15, 33–35].

The assessment instrument (HNNE) was developed in a high-middle-income country (HMIC) and has been standardised against predominant Caucasian populations [36, 37]. Discrepancies in optimality scores were demonstrated between low-risk term-born infants in low-resource settings in Uganda and Ghana and the low-risk term-born infant population in the United Kingdom (UK) where the optimality scoring system was originally developed [38, 39].

Optimality scores based on studies in some developing countries raised questions about the applicability of the optimality scores of the original UK population in such countries [38–41]. Nevertheless, the test continued to successfully indicate deviating performance in high-risk neonatal populations [42–44, 46]. Valuable data were reported on neurodevelopmental screening in various developing countries and resource-limited conditions [38–41, 46, 52].

Further studies were recommended for infants from varying populations whose development can be longitudinally monitored to establish reliable normative global data for this age range [26, 32, 39, 52]. Population-level markers, such as optimality scores, for infants in the age groups 6- and 10 weeks from developing countries may enable health care professionals’ effective monitoring of infant neurodevelopment during this window period, making timeous intervention at an early stage possible [22, 23, 25, 46].

Therefore, the first aim of this study was to assess and attribute scores to the distribution of neuro-behavioural characteristics of healthy 6- and 10-week term infants born from mothers with low-risk pregnancies according to SA ante-natal guidelines [47] in the Tshwane District in SA by implementing the HNNE. The second aim was to apply the HNNE optimality scoring system to the raw scores (RSs) obtained by this cohort of 6- and 10-week term infants. To our knowledge, no data of its kind in this age group has previously been documented.

## Material and Methods

A prospective, longitudinal descriptive study was undertaken to assess and attribute RSs to the distribution of the neuro-behavioural characteristics of 6- and 10-week-old infants in Tshwane District, SA between October 2019 and June 2020. Secondly, the HNNE optimality scoring system was applied to convert the frequency distribution of the RSs to optimality scores.

### Study population

A cohort of thirty-five (35) infants that formed part of an ongoing ante-, peri-, and postnatal investigation - i.e., Umbiflow International Study and the UmbiBaby Study - were selected for participation in this study. Permissions to conduct the study were obtained through the University of Pretoria Research Ethics Committee (reference no. 283/2019) as well as institutional permissions from the relevant health services. Only thirty-five of 81 term-born infants participating in the UmbiBaby Study (19 males; 16 females) who attended both their 6- and 10-week neurological assessments were included in this part of the study [48,49].

The infants were born to mothers who were prospectively enrolled in the Umbiflow Study that screened an unselected population of women with low-risk pregnancies (according to SA antenatal guidelines) [48]. The women were screened in the third trimester of their pregnancy (between 28 and 34 weeks) using the low-cost continuous-wave Doppler ultrasound device (Umbiflow™), developed by the South African Medical Research Council (SAMRC) and Council for Scientific and Industrial Research (CSIR) [49, 50].

For the infant follow-up study mothers in Tshwane District were approached around the time of birth at Pretoria West Hospital, Laudium Community Health Centre, and Kalafong Provincial Tertiary Hospital. The mothers of the infants in this study gave written informed consent to participate in the UmbiBaby Study as a mother-infant pair at the 6-week postnatal clinic visit. The mother-infant pair was followed up at the University of Pretoria’s Research Centre for Maternal, Foetal, Newborn and Child Health Care Strategies at Kalafong Hospital for their 6- and 10-week routine postnatal clinic visits.

The inclusion criteria were infants: 1) from mothers with singleton pregnancies; 2) born to mothers with low-risk pregnancies according to local antenatal care guidelines; 3) with known gestational ages, calculated during the Umbiflow Study; 4) who attended both 6- and 10-week clinic visits. The exclusion criteria were infants born to mothers: 1) younger than 18 years of age; 2) infants with chromosomal, structural abnormalities or severe medical conditions.

The neurological assessments were administered by scoring the neuro-behavioural characteristics of the 35 healthy term-born infants at 6- and 10-weeks postnatal age who attended both follow-up assessments. Demographic and nutritional information of the mother-infant pairs were also collected at every visit as part of the ongoing UmbiBaby investigation. The HNNE was administered by a single qualified physiotherapist experienced in administering and interpreting the assessment tool. The therapist is a presenter of the post-basic continuing professional development course called *Exploring Infant Behaviour*. This course aims at training health care professionals to administer and interpret the HNNE based on infant behaviour, as described by Brazelton and Nugent, (1995) in the Neonatal Behavioural Assessment Scale (NBAS) [50].

### Assessment instrument

The HNNE is a standardised assessment instrument developed to assess the neurological state of newborn term infants in the first 48 hours after birth [51]. The assessment was developed in the UK and has subsequently been used in clinical and research settings for nearly 40 years [30, 31]. The assessment proforma was modified to its current form and the *‘optimality scoring system’* was added and validated on 224 low-risk term infants in the UK within 48 hours after birth [30, 36].

To date, optimality scores have been determined and published for preterm and late preterm infant populations at term-equivalent age [33–36]. Normative data and subsequently optimality scores have not yet been published for term infants beyond this age. In conjunction with neuroimaging and neurophysiological techniques, the HNNE discriminated between clinical patterns associated with neurological conditions such as hypoxic ischaemic encephalopathy, periventricular leukomalacia and intra-ventricular haemorrhage [30, 43, 45]. The HNNE’s correlation with other instruments showed total factor scores of F92.42 = 8.7; p < 0.001; R2 = 0.71 [37, 52]. This examination tool is suitable for clinical application with an administration time of 10 – 15 minutes and has interrater reliability of 96% [31, 40, 53, 54]. The instrument does not require formal training or certification and can be self-taught through the Hammersmith Neurological Examination website [54].

This examination consists of 34 items that are characteristic of the functional state of the newborn infant’s central nervous system. These characteristics are categorised in six domains known as *compounds*: 1) *posture and tone, 2) tone patterns, 3) reflexes, 4) spontaneous movements, 5) abnormal neurological signs*, and *6) orientation and behaviour* [30, 31].

Raw Scores: The items are scored in a standard proforma according to a five-point scale. The description of different responses and characteristics for each item are distributed horizontally over five columns. The column that represents the predominant behaviour of the infant for each item, is circled and scored according to the number of columns 1 to 5. Half-scores are appropriate if the infant’s performance falls between the description of two columns (e.g., a score of 2.5 if the infant displays characteristics between that of columns 2 and 3). These scores are defined as RSs. The RSs are not linear and the *‘optimal score’* for each of the 34 items depended on the frequency distribution of RSs per item for the original study population. For example, for the posture item, a score of 1 to 2 would indicate decreased muscle tone, a score of 3 and 4 would be normal tone and a score of 5 would indicate increased muscle tone [30]. The RS for each item was converted to an ‘*optimality score*’, using the 5th and 10th centiles as cut-off points [30, 36]. Items falling above the 10th centile are given a score of 1 (*optimal*), between the 5th and 10th a score of 0.5 (*borderline*), and below the 5th centile, a score of 0 (*sub-optimal*). For the infants at 6- and 10-weeks post-term age in this study population, the same ‘*optimality scoring system’* was applied to their individual RSs [30, 36].

Compound Scores: Compound scores were calculated for each of the 6 domains: *posture and tone, tone patterns, reflexes, spontaneous movements, abnormal neurological signs*, and *orientation and behaviour*. The compound optimality score for each domain is the sum of the optimality scores for the individual items in the particular domain. This score may range from 0 (if all the items in the compound are *sub-optimal*) to a maximum score equal to the number of items in the particular compound if every item in the domain is *optimal* with a score of 1 [30, 36].

Total score: The total optimality score for the whole examination is the sum of the optimality scores of the 34 individual items. This score may range from 0 (if all the items are *sub-optimal*) to a maximum of 34 (if all the items are *optimal*). The distribution of the compound scores and total optimality scores were described as *optimal* when they fell above the 10th centile, *borderline* between the 5th and 10th centile and *sub-optimal* below the 5th centile [30, 36].

### Clinical Examination

The environmental conditions in which the assessment was performed were controlled for noise, bright light, and temperature. Because the assessment elicited variable behavioural responses in the infants, it was performed with all infants in a baseline behavioural state 4 (quiet alert state) [50]. The assessment was also done halfway between feeds to ensure comparable behaviour amongst infants as far as possible. The infants were examined on a flat wooden table with a thin mat for comfort but assuring a firm and comfortable surface for bony areas, e.g., the occiput during rolling of the head from side to side. After uncovering the infant (only the diaper was left on), the evaluation started with a short period (two minutes) of observing posture and spontaneous movement in the supine position. The HNNE physically challenges infants and therefore the characteristics of the items such as *posture and tone* of the head, trunk and limbs in supine, and the *quality and quantity of spontaneous movement* were assessed before physical and tactile input from the examiner that formed an essential part of the rest of the assessment.

To ensure optimal performance of the infant in a quiet state, the items for *eye appearance, auditory, visual orientation and alertness* in the *orientation and behaviour* domains were recorded following the assessment of *posture* and *spontaneous movement*. This sequence in performing the assessment enabled the examiner to successfully complete the assessment, noting the triggers in the infant’s *irritability* and *crying* behaviour. The *Moro reflex* was administered at the end of the assessment as infants are often startled by their own response and may then be difficult to console. The assessment ended with the measurement of the infant’s *head circumference*.

The findings for each item in the domains of *posture and tone, tone patterns, reflexes, spontaneous movements, abnormal neurological signs*, and *orientation and behaviour* were recorded during the examination. The duration of the assessment was between 10 - 15 minutes but depended on the infants’ behavioural state and responses. The infants’ *consolability* (strategies required to be consoled), the intensity of the *cry* and general *irritability* (response to the handling and physical challenges of the examination) were assessed throughout the examination and were recorded last.

## Results

The bio-demographic data of the participating mother-infant pairs in this study are shown in Table 1. The infants resided in formal (60%) and informal settlements (40%) in the Tshwane District. The majority (60%) were delivered by Caesarean section and 40% were delivered through normal vaginal delivery. The mean gestational age- and sex-normalised z-scores for weight, length and head circumference were assessed at birth using the INTERGROWTH – 21st Newborn Size at Birth standard [55]. Age- and sex-normalised and weight, length, head circumference and weight-for-length z-scores were assessed at 6 and 10 weeks using the WHO child growth standards [56]. The mean z-scores for infant anthropometry at birth, 6 and 10 weeks all lay between −1 and +1, indicating that the majority of the infants fell within the range for optimal growth (i.e. between the z-scores of −2 and +2). The average chronological ages at the follow-up assessments (i.e., mean time since birth) were 6.4 and 10.4 weeks, respectively. Twenty-five percent (25.7%) of mothers were HIV-infected, however, none of the infants in the study tested HIV positive.

**Table 1.**
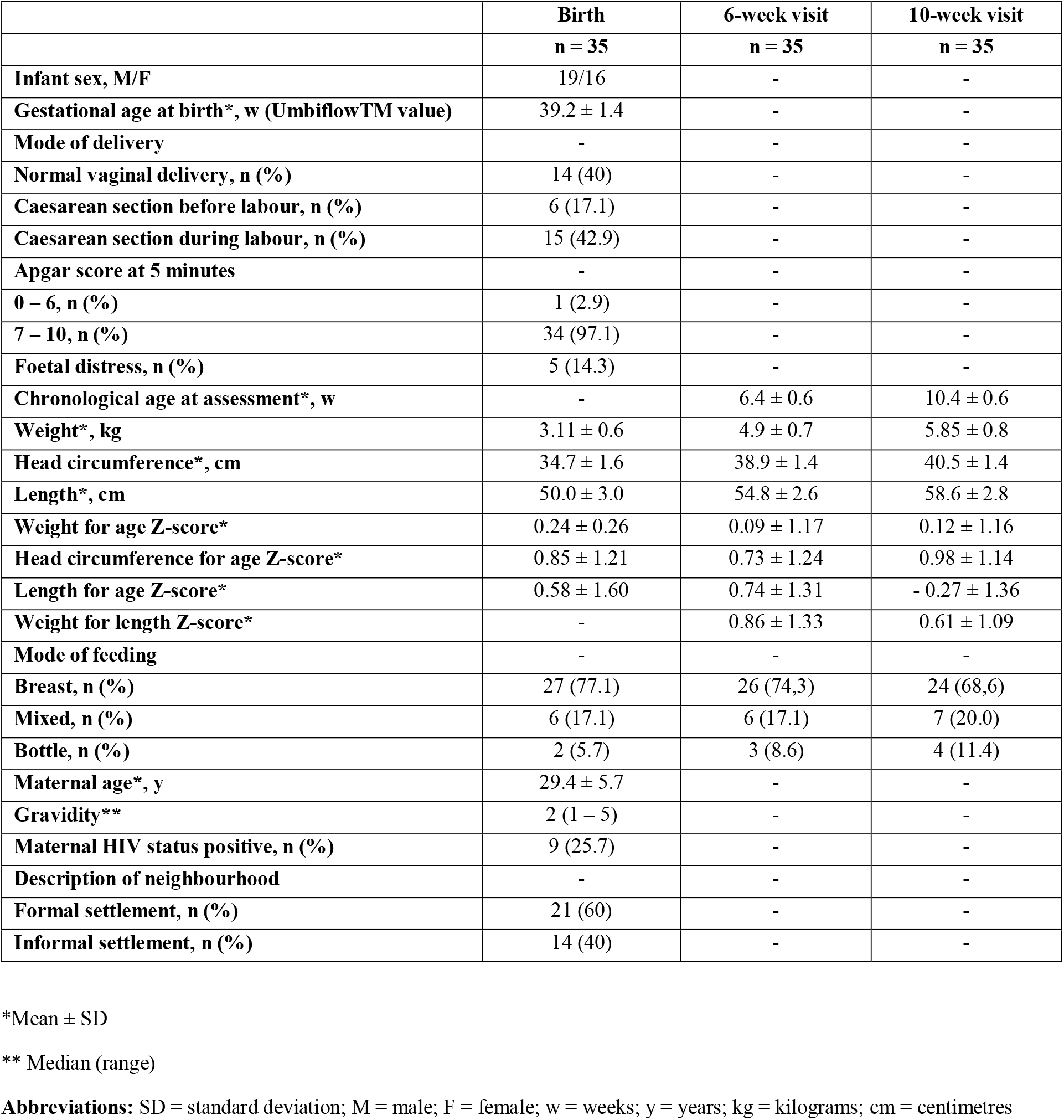
Bio-demographic data of the mother-infant pairs in this study population.

|  | **Birth** | **6-week visit** | **10-week visit** |
| --- | --- | --- | --- |
|  | **n = 35** | **n = 35** | **n = 35** |
| **Infant sex, M/F** | 19/16 | - | - |
| **Gestational age at birth\*, w (Umbiflow™ value)** | 39.2 ± 1.4 | - | - |
| **Mode of delivery** | - | - | - |
| **Normal vaginal delivery, n (%)** | 14 (40) | - | - |
| **Caesarean section before labour, n (%)** | 6 (17.1) | - | - |
| **Caesarean section during labour, n (%)** | 15 (42.9) | - | - |
| **Apgar score at 5 minutes** | - | - | - |
| **0 – 6, n (%)** | 1 (2.9) | - | - |
| **7 – 10, n (%)** | 34 (97.1) | - | - |
| **Foetal distress, n (%)** | 5 (14.3) | - | - |
| **Chronological age at assessment\*, w** | - | 6.4 ± 0.6 | 10.4 ± 0.6 |
| **Weight\*, kg** | 3.11 ± 0.6 | 4.9 ± 0.7 | 5.85 ± 0.8 |
| **Head circumference\*, cm** | 34.7 ± 1.6 | 38.9 ± 1.4 | 40.5 ± 1.4 |
| **Length\*, cm** | 50.0 ± 3.0 | 54.8 ± 2.6 | 58.6 ± 2.8 |
| **Weight for age Z-score\*** | 0.24 ± 0.26 | 0.09 ± 1.17 | 0.12 ± 1.16 |
| **Head circumference for age Z-score\*** | 0.85 ± 1.21 | 0.73 ± 1.24 | 0.98 ± 1.14 |
| **Length for age Z-score\*** | 0.58 ± 1.60 | 0.74 ± 1.31 | - 0.27 ± 1.36 |
| **Weight for length Z-score\*** | - | 0.86 ± 1.33 | 0.61 ± 1.09 |
| **Mode of feeding** | - | - | - |
| **Breast, n (%)** | 27 (77.1) | 26 (74.3) | 24 (68.6) |
| **Mixed, n (%)** | 6 (17.1) | 6 (17.1) | 7 (20.0) |
| **Bottle, n (%)** | 2 (5.7) | 3 (8.6) | 4 (11.4) |
| **Maternal age\*, y** | 29.4 ± 5.7 | - | - |
| **Gravidity\*\*** | 2 (1 – 5) | - | - |
| **Maternal HIV status positive, n (%)** | 9 (25.7) | - | - |
| **Description of neighbourhood** | - | - | - |
| **Formal settlement, n (%)** | 21 (60) | - | - |
| **Informal settlement, n (%)** | 14 (40) | - | - |

\*Mean ± SD
\*\* Median (range)
**Abbreviations:** SD = standard deviation; M = male; F = female; w = weeks; y = years; kg = kilograms; cm = centimetres

### Clinical examination results

The 34 items of the HNNE proforma are illustrated in Figures 1 to 5, reflecting the 6 *compounds* of the examination [30, 36]. The distribution of RSs for all items at 6 and 10 weeks respectively and the percentage of infants who scored each RS are illustrated by a small table on the right side of every item (Figs. 1 – 5). The distribution of RSs was determined by the frequency in which the characteristics were displayed in the performance of the infant in each of the 34 items. The scores that were frequently obtained by infants in both age groups and that were considered *optimal* (above the 10th centile) by a score of 1 are indicated in the dark-grey areas. The light-grey columns indicate those RSs that were considered *borderline* (between the 5th – 10th centile) by a score of 0.5. The RSs that fell outside the grey areas are considered *sub- optimal* (below the 5th centile) and therefore scored 0.

**Figure 1.**
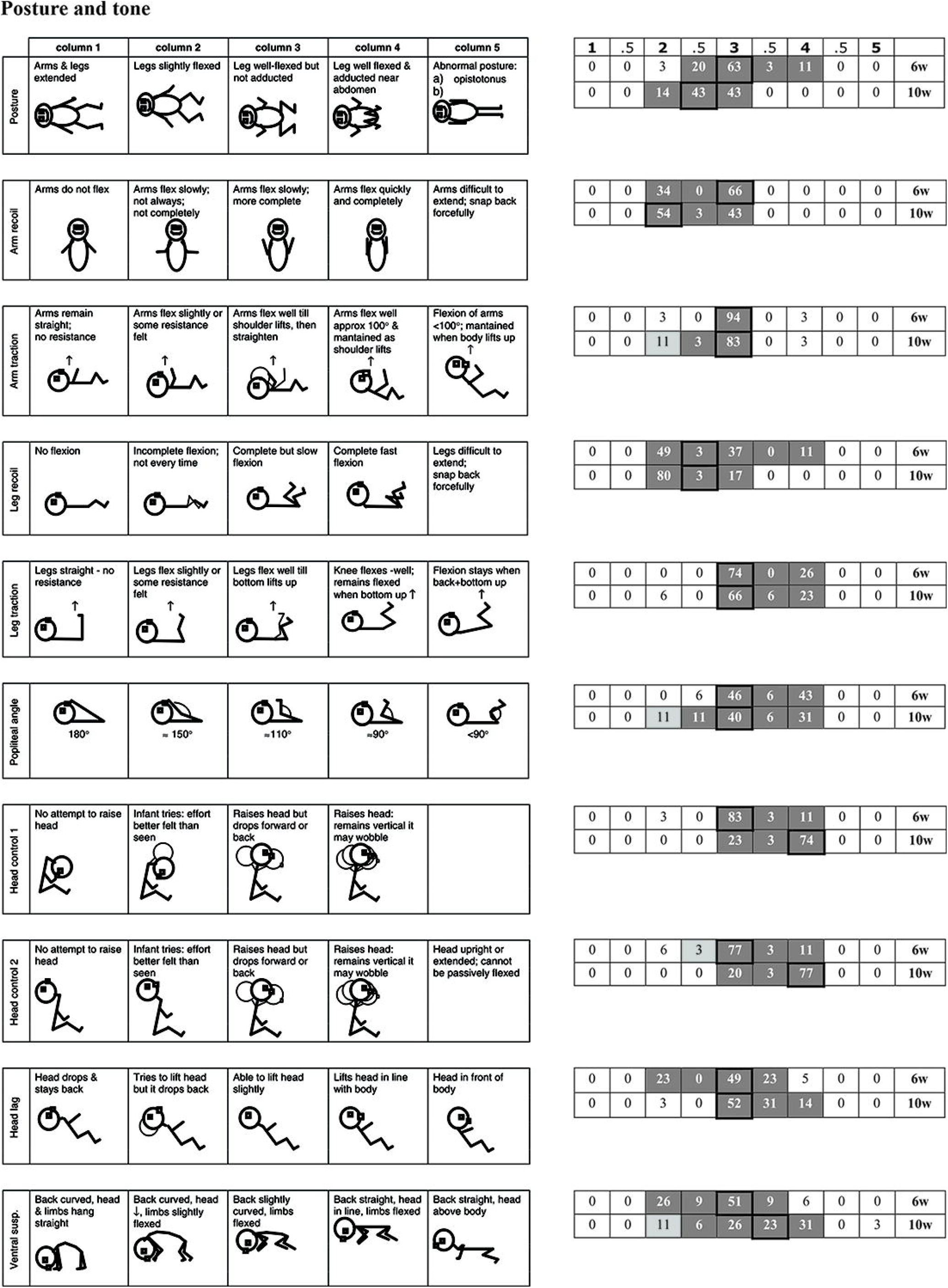
Raw score distribution of *posture and tone* items. Scores that were frequently obtained by infants in each age group and considered optimal (≥ 10th centile) by a score of 1, are indicated in the dark-grey areas in the small tables on the right of the HNNE proforma [30]. The light-grey columns indicate RSs considered borderline (5th – 10th centile) by a score of 0.5. Sub-optimal RSs (< 5^th^ centile) that fell outside the grey areas, scored 0. The cell with the highlighted border represents the median score. The same *‘optimality scoring’* principle applies to Figures 1 – 5 [30, 36].

Compound optimality (summation of individual item optimality scores) and the total optimality score (summation of the 34 individual item optimality scores) ranges, mean values, standard deviations and percentages for the whole population at 6- and 10-weeks postnatal age are shown in Table 2.

**Table 2:** Compound and total optimality scores for healthy term infants at 6- and 10- weeks post-term age (n = 35)

| *Compounds* | Age (w*) | Range | Mean | SD | 5th | 10th | 90th | 95th | Optimal score (%) |
| --- | --- | --- | --- | --- | --- | --- | --- | --- | --- |
| **1. Posture & tone** | 6 | 8 – 10 | 9.67 | 0.58 | 8.7 | 9 | 10 | 10 | 97 |
|  | 10 | 8 – 10 | 9.71 | 0.52 | 8.7 | 9.2 | 10 | 10 | 94 |
| **2. Tone patterns** | 6 | 4 – 5 | 4.87 | 0.31 | 4 | 4.5 | 5 | 5 | 97 |
|  | 10 | 4 – 5 | 4.97 | 0.17 | 5 | 5 | 5 | 5 | 97 |
| **3. Reflexes** | 6 | 3 – 6 | 5.64 | 0.72 | 4 | 4.7 | 6 | 6 | 86 |
|  | 10 | 3 – 6 | 5.80 | 0.62 | 4.7 | 5.5 | 6 | 6 | 94 |
| **4. Spontaneous movements** | 6 | 1.5 – 3 | 2.87 | 0.35 | 2 | 2.5 | 3 | 3 | 94 |
|  | 10 | 1 – 3 | 2.83 | 0.51 | 1.7 | 2.4 | 3 | 3 | 86 |
| **5. Abnormal signs** | 6 | 1 – 3 | 2.86 | 0.43 | 2 | 2.4 | 3 | 3 | 86 |
|  | 10 | 2 – 3 | 2.89 | 0.32 | 2 | 2.4 | 3 | 3 | 86 |
| **6. Orientation & behaviour** | 6 | 3 – 7 | 6.54 | 1.09 | 3.7 | 5.4 | 7 | 7 | 86 |
|  | 10 | 5 – 7 | 6.74 | 0.51 | 5.9 | 6 | 7 | 7 | 100 |
| **Total** | 6 | 25.5 – 34 | 32.46 | 2.02 | 28.9 | 29.5 | 34 | 34 | 91 |
|  | 10 | 29 – 34 | 32.94 | 1.20 | 30.9 | 31.5 | 34 | 34 | 94 |

\*w = weeks postnatal age

#### Posture and tone

The frequency distribution of RSs in this domain was different for each of the 10 items at both 6 and 10 weeks (Fig. 1). Median score shifts of one and two columns to the left from 6 weeks to 10 weeks indicated characteristics of diminished flexion posture and lesser resistance of upper limbs to traction and passive extension respectively. Median score shifts of one to two columns involving flexor and extensor head control in sitting and extensor control in horizontal occurred to the right from 6 weeks to 10 weeks. The compound optimality scores for *posture and tone* ranged from 8 – 10 out of 10 at both 6 and 10 weeks. These scores were achieved by 97% of the total population of infants at the 6-week and 94% at the 10-week postnatal age. Scores of 8.5 and lower at both 6 and 10 weeks were considered sub-optimal (Table 2).

#### Tone patterns

Scores in this domain of the HNNE were obtained by comparing the RSs of selected individual items of limb tone and head control patterns from the *tone and posture* compound (Fig. 2). At 6 weeks 97% of infants scored ≥ 4.5 out of 5 and at 10 weeks 97% scored 5 out of 5. The distribution and median scores at 6 and 10 weeks were consistent in all 5 items. (Table 2).

**Figure 2.**
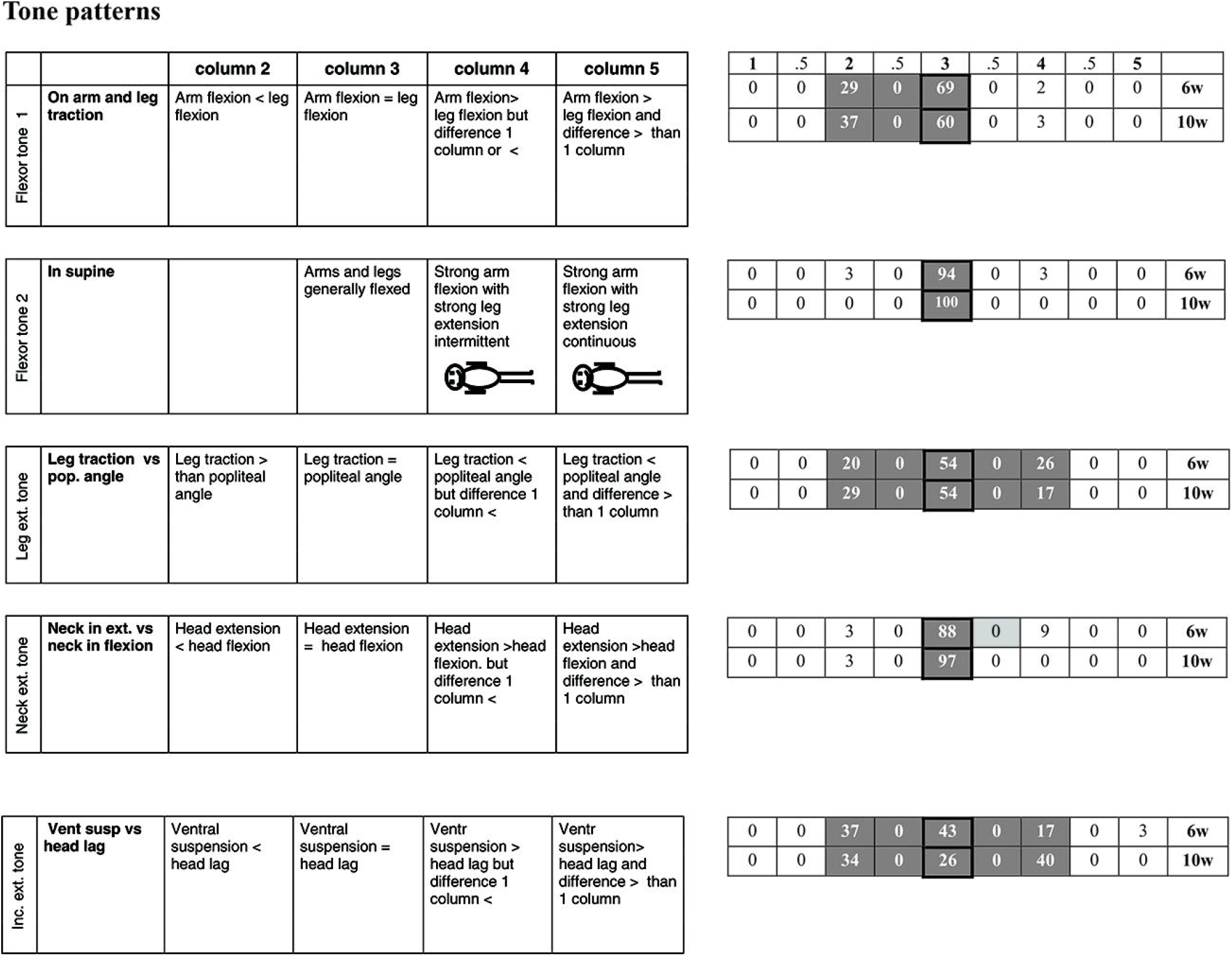
Raw score distribution of *tone pattern* items. See score description under Fig.1.

#### Reflexes

Palmar grasp RSs at 10 weeks were more widely distributed and showed a median score shift of two columns to the left on the proforma examination form indicating a diminished reflex response towards 10 weeks (Fig. 3). The compound optimality score for *reflexes* ranged between 3 – 6 out of 6 in both the 6 and 10-week age groups. Scores from 5 to 6 were found in 86% of infants at 6 weeks and in 94% of infants at 10 weeks. (Table 2).

**Figure 3.**
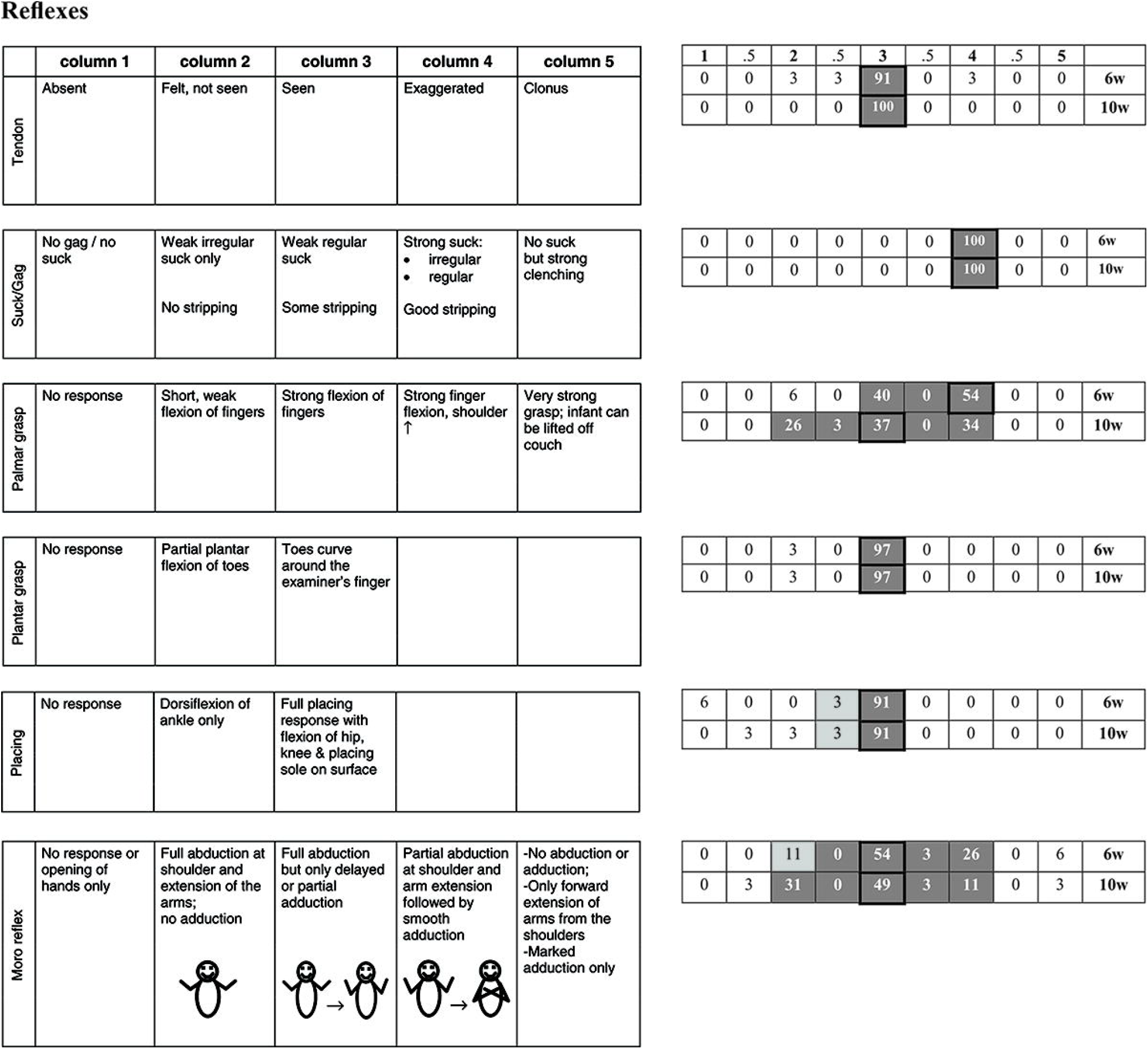
Raw score distribution of *reflex* items. See score description under Fig.1.

#### Movements

The *head in prone* item at 10 weeks displayed a wider distribution towards the right from the mutual median RS. This distribution is indicative of increasing postural extensor tone in horizontal. The compound optimality score for *movements* ranged from 1.5 to 3 out of 3 at 6 weeks and 1 to 3 out of 3 at 10 weeks (Fig. 4). The optimal score of ≥ 2.5 was achieved by 94% of infants at 6 weeks and 86% of infants at 10 weeks. (Table 2).

**Figure 4.**
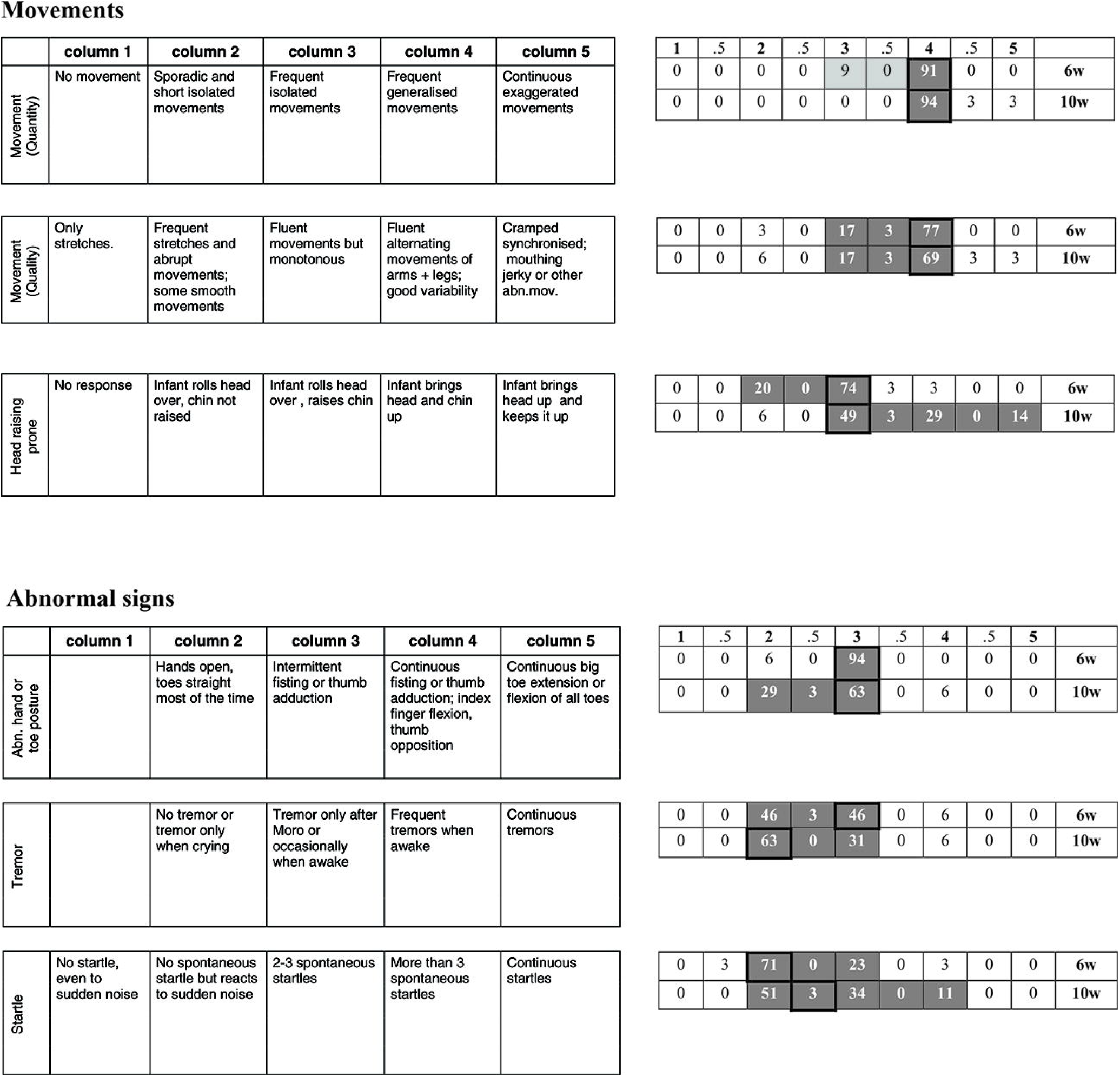
Raw score distribution of *movement* and *abnormal signs* items. See score description under Fig.1.

#### Abnormal signs

The median RS for tremors at 10 weeks shifted two columns towards the left from the 6-week median score, indicating lesser tremor behaviour. The median RS for startle behaviour at 10 weeks demonstrated a one column shift to the right from the 6-week median score (Fig. 4). The compound optimality scores for *abnormal signs* ranged between 1 and 3 out of 3 at 6 weeks and 1.5 and 3 out of 3 at 10 weeks. A compound optimality score of 2.5 was achieved by 86% of the 6-week-old infants and a score of 2.5 by 86% of the 10-week-old infants (Table 2).

#### Orientation & behaviour

For both items involving *visual orientation* and *alertness* at 10 weeks, the median RSs shifted to the right with two columns from the 6-week median score. The median RS for *consolability* at 10 weeks shifted one column to the left from the 6-week median score (Fig. 5). The compound optimality scores for *orientation and behaviour* ranged from 3 to 7 out of 7 at 6 weeks and from 5 to 7 out of 7 at 10 weeks (Fig. 6). Scores of 5 to 7 were found in 86% of infants at 6 weeks and scores of 6 and 7 in 100% of infants at 10 weeks (Table 2).

**Figure 5.**
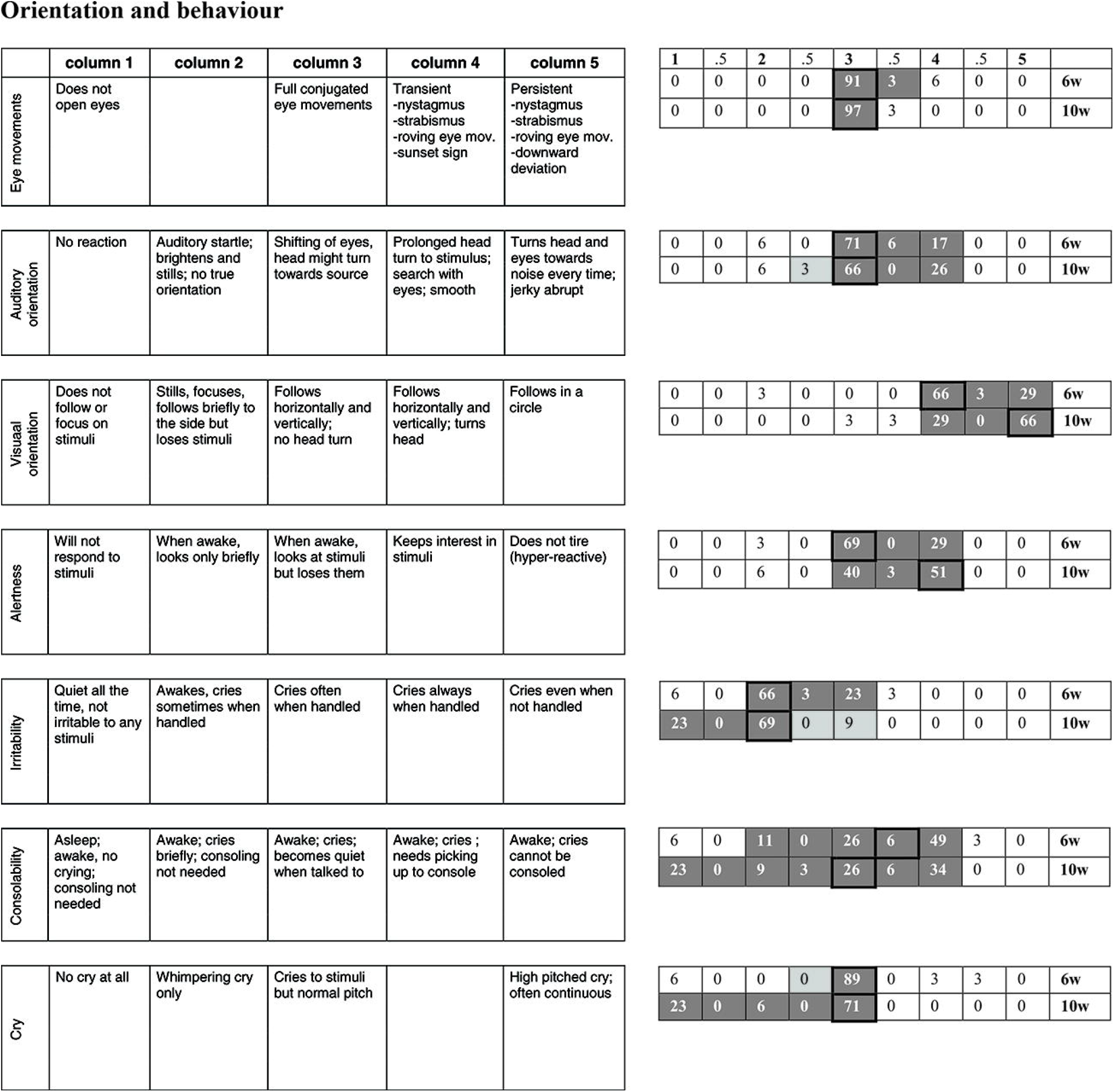
Raw score distribution of *orientation and behaviour* items. See score description under Fig.1.

#### Total examination optimality score

A total optimality score for the examination was obtained when the optimal scores of all 34 items were added up. In this study, the total optimality scores for infants at 6-weeks post-term age ranged between 25.5 and 34 out of the 34 items. The range for total optimality scores of infants at 10-weeks post-term age was 29 to 34 out of 34 items. In the 6-week age group, 91% of the infants obtained a total examination optimality score between 29 and 34. Scores ≤ 28.5 at 6 weeks were considered suboptimal. In the 10-week age group, 94% of the infants obtained total examination optimality scores between 31.5 and 34. A score ≤ 30.5 at 10 weeks was considered sub-optimal (Table 2).

## Discussion

Despite the progress in research, programmes and policies that promote the health and well-being of mothers and children, 66% of children younger than five (5) years in developing countries still risk failing to reach their developmental potential [16, 17]. Many maternal and foetal conditions are associated with adverse insults to the developing brain that cause altered brain structure and sub-optimal neurodevelopmental trajectories in the affected infant [1, 10, 13, 20].

Health care professionals are urgently called to identify at-risk infants but also contribute to sustainable early childhood intervention by addressing and thus potentially limiting the risk of adverse short- and long-term development outcomes in surviving and high-risk infants [16, 19, 22, 39]. The existing studies that examined the neurological status and developmental characteristics of newborn infants, published normative data with optimality scores for term and late preterm infants as well as preterm infants at term equivalent age [33–36].

Although these scores are very useful in a variety of settings, questions were raised about the validity of the original optimality scores of infants in settings other than developed countries [38–41, 52]. Guzzetta et al., (2005) are the only researchers that studied infants longitudinally in the same age groups that compare to our study. These authors’ randomly studied 79 low-risk infants from birth to 10 weeks. The study provided useful guidelines for screening infants at 6 weeks, however, values such as optimality scores that can be used in well-baby clinics to identify infants at risk of developmental delay post-natal age were not provided due to the small numbers of infants per post-natal age group. Their findings prompted us to use the HNNE in our longitudinal study and apply the ‘*optimality scoring system*’ to a population of healthy infants aged 6- and 10 weeks in a developing country.

Our study assessed and documented the neuro-motor and neuro-behavioural characteristics that were displayed by 35 healthy term-born infants at the ages of 6 and 10 weeks during scheduled clinic visits in a developing country. The RSs obtained on the HNNE in this cohort of infants were converted to optimality scores by applying the HNNE optimality scoring system with cut- off points that resulted in quantitative estimation of each item on the evaluation form (Supplementary Tables S1 - 6) [36].

The results of this study are similar to the observations made by Guzetta et al., (2005) at 6 weeks and support 6 weeks as an important milestone in the neurodevelopmental behaviour in infants in the domains of *posture, limb/axial tone*, and *visual behaviour* [26]. Our study was able to quantify the results per 6- and 10- week post-natal age groups respectively due to higher numbers (n = 35) of participants per age group. Furthermore, the results of this study showed the direction of notable ongoing changes in *upper* and *lower limb tone, active head control* in horizontal and vertical positions, and advanced *visual orientation and alertness* in this cohort of infants as they developed towards 10 weeks. These changes in characteristics indicate that 10 weeks is another significant milestone in the neurodevelopmental process. This trajectory of development has not been described or reported in any of the consulted literature.

The results show different RS distributions for most items in the HNNE proforma in each of the two age groups, as well as different compound and total optimality score ranges respectively. The strength of the study lies in the longitudinal monitoring of the developmental trajectory of milestone characteristics between 6 and 10 weeks’ post-term age in a cohort of healthy infants in the Tshwane District of SA. These characteristics appear to be important changes that occur in healthy term infants across cultures and are therefore considered an important stepping stone for the early identification of neurodevelopmental risk factors in infants at 6 and 10 weeks.

## Conclusion

This study presents the first results of ongoing research and evolving data for identifying neuro- motor and neuro-behavioural characteristics of healthy term-born 6- and 10-week-old infants in a developing country. Studying bigger cohorts in SA may result in the collection of data that can be generalised and then incorporated into the RtHB. Quantifying the observation and grading of the characteristics of the infants’ neuro-motor and neuro-behavioural responses at 6 and 10 weeks respectively, will enable health care professionals to red-flag infants at risk of neurodevelopmental pathology or delay. Such data will lead to a greater understanding, continuous monitoring and effective management of infants and as such contribute to achieving Child Health Sustainable Development Goals in SA.

## Data Availability

All relevant data are within the manuscript and its Supporting Information files.

## Acknowledgements

The authors pay tribute to the mothers and infants who took part in this study. Their contribution provides invaluable information for interpreting infant development in South Africa. Our gratitude is extended to the research team for their administrative role in the recruitment, anthropometry, and the collecting of background information. In her private capacity, the first author wishes to express a sincere appreciation toward the Department of Paediatrics and the University of Pretoria’s Research Centre for Maternal, Foetal, Newborn and Child Health Care Strategies at Kalafong Provincial Tertiary Hospital for the opportunity to participate in the UmbiBaby Study.

## Author contributions

**Conceptualization:** Marna Nel, Carina Eksteen, Ute Feucht

**Data curation:** Marna Nel, Helen Mulol

**Formal analysis:** Marna Nel, Helen Mulol

**Investigation:** Marna Nel

**Methodology:** Marna Nel, Carina Eksteen

**Project administration:** Marna Nel, Ute Feucht, Helen Mulol

**Resources:** Ute Feucht, Helen Mulol, Marna Nel

**Supervision:** Ute Feucht, Helen Mulol, Carina Eksteen

**Validation:** Ute Feucht, Marna Nel, Carina Eksteen

**Visualization:** Marna Nel, Carina Eksteen

**Writing – original draft:** Marna Nel

**Writing – review & editing:** Marna Nel, Ute Feucht, Helen Mulol, Carina Eksteen

## Supporting information

**S1 Table. Conversion of raw scores into optimality scores for items assessing *posture and tone***.

(DOCX)

**S2 Table. Conversion of raw scores into optimality scores for *tone patterns***.

(DOCX)

**S3 Table. Conversion of raw scores into optimality scores for items assessing *reflexes***.

(DOCX)

**S4 Table. Conversion of raw scores into optimality scores for items assessing *movements***.

(DOCX)

**S5 Table. Conversion of raw scores into optimality scores for items assessing *abnormal signs***.

(DOCX)

**S6 Table. Conversion of raw scores into optimality scores for items assessing *orientation and behaviour***.

(DOCX)

