## Supplementary Tables 1 - 6 for "Hammersmith Neonatal Neurological Examination of healthy term infants at ages 6 and 10 weeks in Tshwane District, South Africa"

**S1 Table. Conversion of raw scores into optimality scores for items assessing *posture and tone*.**

| **Posture & tone** | **Scores at 6 weeks** | | | **Scores at 10 weeks** | | |
| --- | --- | --- | --- | --- | --- | --- |
|  | **1** | **0.5** | **0** | **1** | **0.5** | **0** |
| **Posture** | 2.5;3;4 |  | <2.5;>4 | 2;3 |  | <2;>3 |
| **Arm recoil** | 2;3 |  | <2;>3 | 2;3 |  | <2;>3 |
| **Arm traction** | 3 |  | <3;>3 | 2.5;3 | 2 | <2;>3 |
| **Leg recoil** | 2;3;4 |  | <2;>4 | 2;3 |  | <2;>3 |
| **Leg traction** | 3;4 |  | <3;>4 | 3;4 |  | ≤2.5;>4 |
| **Popliteal angle** | 3;4 |  | ≤2.5;>4 | 2.5;3;4 | 2 | <2;>4 |
| **Head control 1** | 3;4 |  | <3;>4 | 3;4 |  | <3;>4 |
| **Head control 2** | 3;4 | 2.5 | <2.5;>4 | 3;4 |  | <3;>4 |
| **Head lag** | 2;3.5 |  | <2;≥4 | 3;4 |  | <3;>4 |
| **Ventral suspension** | 2;3.5 |  | <2;≥4 | 2.5;3;4 | 2 | <2;>4 |

A score of 1 (≥10th centile) was given to the raw scores found in 90% of the population or more, a score of 0.5 (5th-10th percentile) to those found in more than 5% but less than 10%, and a score of 0 (<5th centile) to those found in less than 5%. Note that the optimality scores change with increasing postnatal age [30, 31].

**S2 Table. Conversion of raw scores into optimality scores for *tone patterns*.**

| **Tone patterns** | **Scores at 6 weeks** | | | **Scores at 10 weeks** | | |
| --- | --- | --- | --- | --- | --- | --- |
|  | **1** | **0.5** | **0** | **1** | **0.5** | **0** |
| **Flexor tone arm & leg traction** | 2;3 |  | <2;>3 | 2;3 |  | <2;>3 |
| **Flexor tone posture (supine)** | 3 |  | <3;>3 | 3 |  | <3;>3 |
| **Leg extensor tone** | 2;3;4 |  | <2;>4 | 2;3;4 |  | <2;>4 |
| **Head control 1&2** | 3 | 4 | <3;>4 | 3 |  | <3;>3 |
| **Head lag & ventral suspension** | 2;3;4 |  | <2;>4 | 2;3;4 |  | <2;>4 |

Scoring system as in S1 Table.

**S3 Table. Conversion of raw scores into optimality scores for items assessing *reflexes*.**

| **Reflexes** | **Scores at 6 weeks** | | | **Scores at 10 weeks** | | |
| --- | --- | --- | --- | --- | --- | --- |
|  | **1** | **0.5** | **0** | **1** | **0.5** | **0** |
| **Tendon** | 3 |  | ≤2.5;>3 | 3 |  | <3;>3 |
| **Sucking** | 4 |  | <4;>4 | 4 |  | <4;>4 |
| **Palmar grasp** | 3;4 |  | ≤2.5;>4 | 2;3;4 |  | <2;>4 |
| **Plantar grasp** | 3 |  | <3 | 3 |  | <3 |
| **Placing** | 3 | 2.5 | ≤2 | 3 | 2.5 | ≤2 |
| **Moro reflex** | 2.5;3;4 | 2 | <2;>4 | 2;3;4 |  | <2;>4 |

Scoring system as in S1 Table.

**S4 Table. Conversion of raw scores into optimality scores for items assessing *movements*.**

| **Spontaneous movement** | **Scores at 6 weeks** | | | **Scores at 10 weeks** | | |
| --- | --- | --- | --- | --- | --- | --- |
|  | **1** | **0.5** | **0** | **1** | **0.5** | **0** |
| **Quantity** | 4 | 3;3.5 | <3;>4 | 4 |  | <4;>4 |
| **Quality** | 3;4 |  | <3;>4 | 3;4 |  | ≤2.5;>4 |
| **Head raising** | 2;3 |  | <2;≥3.5 | 3;4;5 |  | ≤2.5 |

Scoring system as in S1 Table.

**S5 Table. Conversion of raw scores into optimality scores for items assessing *abnormal signs*.**

| **Abnormal signs** | **Scores at 6 weeks** | | | **Scores at 10 weeks** | | |
| --- | --- | --- | --- | --- | --- | --- |
|  | **1** | **0.5** | **0** | **1** | **0.5** | **0** |
| **Hand postures** | 3 |  | ≤2.5;>3 | 2;3 |  | <2;≥3.5 |
| **Tremors** | 2;3 |  | <2;≥3.5 | 2;3 |  | <2;≥3.5 |
| **Startles** | 2;3 |  | <2;>3 | 2;3;4 |  | <2;>4 |

Scoring system as in S1 Table.

**S6 Table. Conversion of raw scores into optimality scores for items assessing *orientation and behaviour*.**

| **Behaviour** | **Scores at 6 weeks** | | | **Scores at 10 weeks** | | |
| --- | --- | --- | --- | --- | --- | --- |
|  | **1** | **0.5** | **0** | **1** | **0.5** | **0** |
| **Eyes** | 3;3.5 |  | <3;≥4 | 3 |  | <3;>3 |
| **Auditory orientation** | 3;4 |  | ≤2.5;>4 | 3;4 | 2.5 | ≤2;>4 |
| **Visual orientation** | 4;5 |  | <4 | 4;5 |  | ≤3.5 |
| **Alertness** | 3;4 |  | <3;>4 | 3;4 |  | ≤2.5;>4 |
| **Irritability** | 2;3 |  | <2;>3 | 1;2 | 2.5;3 | >3 |
| **Consolability** | 2;3;4 |  | <2;>4 | 1;2;3;4 |  | >4 |
| **Cry** | 3 | 2.5 | <2;≥3 | 1;2;3 |  | >3 |

Scoring system as in S1 Table.
